# Life course air pollution exposure and cognitive decline: modelled historical air pollution data and the Lothian Birth Cohort 1936

**DOI:** 10.1101/2020.10.16.20163691

**Authors:** Tom C. Russ, Mark P. C. Cherrie, Chris Dibben, Sam Tomlinson, Stefan Reis, Ulrike Dragosits, Massimo Vieno, Rachel Beck, Ed Carnell, Niamh K. Shortt, Graciela Muniz-Terrera, Paul Redmond, Adele M. Taylor, Tom Clemens, Martie van Tongeren, Raymond M Agius, John M. Starr, Ian J. Deary, Jamie R. Pearce

## Abstract

**Background:** Air pollution has been consistently linked with dementia and cognitive decline. However, it is unclear whether risk is accumulated through long-term exposure or whether there are sensitive/critical periods. A key barrier to clarifying this relationship is the dearth of historical air pollution data.

**Objective:** To demonstrate the feasibility of modelling historical air pollution data and using them in epidemiological models.

**Methods:** Using the EMEP4UK atmospheric chemistry transport model, we modelled historical fine particulate matter (PM_2.5_) concentrations for the years 1935, 1950, 1970, 1980, and 1990 and combined these with contemporary modelled data from 2001 to estimate life course exposure in 572 participants in the Lothian Birth Cohort 1936 with lifetime residential history recorded. Linear regression and latent growth models were constructed using cognitive ability (IQ) measured by the Moray House Test at the ages of 11, 70, 76, and 79 years to explore the effects of historical air pollution exposure. Covariates included sex, IQ at age 11 years, social class, and smoking.

**Results:** Higher air pollution modelled for 1935 (when participants would have been *in utero*) was associated with worse change in IQ from age 11-70 years (β=-0.006, SE=0.002, P=0.03) but not cognitive trajectories from age 70-79 years (P>0.05). There was no support for other critical/sensitive periods of exposure or an accumulation of risk (all P>0.05).

**Conclusions:** The life course paradigm is essential in understanding cognitive decline and this is the first study to examine life course air pollution exposure in relation to cognitive health.

## INTRODUCTION

Dementia is a global public health crisis with almost 47 million people affected in 2015 and almost 10 million new cases every year, leading to a projected prevalence of over 130 million by 2050[1]. The brain changes which lead to many dementias – including the most common form, Alzheimer’s dementia – begin in midlife and only manifest in later life[2]. Dementia prevention is now a worldwide priority and accepted risk factors include lower levels of educational attainment (in early life), cardiovascular disease risk factors (with hypertension and obesity particularly highlighted in mid-life), depression, hearing loss, and possession of the APOE ε4 allele[3]. A recent Lancet Commission report and other analyses have estimated that approximately a third of dementia risk can be explained by these common risk factors[4, 5]. With genetic factors (most prominently APOE ε4 carriage) explaining approximately another third[6], this leaves around a third of dementia risk unexplained. However, there is also evidence linking a number of environmental risk factors with dementia which might account for some of this unexplained risk[7]. The risk factor for which there is strongest evidence is air pollution[8, 9]. However, the field has been criticised since studies to date have lacked long-term (i.e. whole life) assessment of both exposure and outcome[10]. Thus, no light has yet been shed on the question of *when* in the life-course exposure to air pollution is most harmful to the brain. Recent papers describing “long-term exposure” to air pollution only estimated air pollution exposure at one time point[11, 12]. Indeed, answering this question from a life-course epidemiology perspective is hampered by both a dearth of available air pollution data from earlier than a few decades ago, when systematic long-term monitoring of atmospheric concentrations was implemented and limited information about the geographical location of study participants over their lives[13]. Therefore, we modelled air pollution data (fine particulate matter, with an aerodynamic diameter of 2.5μm or smaller; PM_2.5_) for multiple time periods and linked these with the Lothian Birth Cohort 1936 (LBC1936) — for whom lifetime residential history is available — to investigate links between air pollution and cognitive change over more than six decades.

## MATERIALS & METHODS

### Study participants

The LBC1936 is a well-established cohort study, originally comprising 1091 men and women aged approximately 70 years at recruitment. Almost all sat the Moray House Test (MHT) of general cognitive ability in the Scottish Mental Survey in 1947 when they were aged about 11 years[14]. Thus, general intelligence data are available for almost all participants at ages 11 years, and repeatedly from approximately 70 years onwards. In the present study, we used data from waves 1, 3 and 4 when participants had mean ages of 69.5 (SD=0.8), 76.3 (0.7), and 79.3 (0.6) years respectively; the MHT was not administered in wave 2. We operationalised cognitive function in the same way as previous studies, adjusting for age in days and standardising to an IQ-type score with mean 100 and SD 15[15]. In line with previous analyses, change in IQ score was computed as the standardised residual from a linear regression model with age 11 IQ as the independent variable and age 70 IQ as the dependent variable; this is superior to computing the arithmetic difference[16, 17].

In 2014, surviving LBC1936 participants were asked to complete a lifetime residential questionnaire and 593 of 704 approached provided usable life grid data (full addresses) which were geocoded to latitude and longitude[15]. Participants had a mean (SD) 11.3 (2.9) separate locations throughout life, ranging from six to 27, with the years they lived there also recorded. Each location was allocated to the closest time period for which air pollution data were available: 1935 (location year 1942 or earlier); 1950 (1943-1959); 1970 (1960-1975); 1980 (1976-1985); 1990 (1986-1995); or 2001 (1995-2004); locations after 2004 were excluded to avoid overlap with cognitive testing (wave 1 of the LBC1936 took place from 2004-2007[14]). Participants may have had more than one location allocated to each time point — e.g., all locations between the years 1995 and 2004 would be allocated to the 2001 time point. Thus, participants had up to ten locations per time point (mean [SD] values ranged from 1.11 [0.34] locations recorded in 2001 to 3.38 [1.33] locations in 1970). Twenty one participants (3.5%) were missing location data for at least one time point, leaving 572 in the final sample who had location (and therefore air pollution) data available for every time point.

Other covariate data available in the LBC1936 and used in the models included sex, parental occupational social class (using the Registrar General 1951 classification from I to V[18]), and self-reported smoking status (current smoker at wave 1 or non-/ex-smoker).

### Air pollution modelling

The EMEP4UK atmospheric chemistry transport model (rv4.3 for 1970-2010 and rv4.10 for 1935/50 [19]) was used to model historical ambient concentrations of fine particulate matter (PM_2.5_) for the years 1935, 1950, 1970, 1980, and 1990 which were combined with contemporary modelled data from 2000 onwards and residential histories to estimate life course exposure. The EMEP4UK model setup, geographical coverage, and configuration used here has been described previously[20, 21]. The model covers the European Union with a horizontal resolution of 0.5° x 0.5° used to provide the boundary condition for a nested UK domain (resolution of 0.055° x 0.055°). The modelled PM_2.5_ and other key air pollutant concentrations are routinely validated against observations across UK monitoring networks[20-25] and have been used for the assessment of population exposure over longer time scales in other studies for the period 1970 to 2010[26]. Emission data have been identified as key sources of uncertainty in modelling historic air pollution. A detailed assessment of sensitivity and uncertainty of the Atmospheric Chemistry Transport Models (ACTM) applied in this study has been published elsewhere[27].

UK-specific gridded emissions of nitrogen oxides (NO_x_), sulphur oxides (SO_x_), ammonia (NH_3_), non-methane volatile organic compounds (NMVOCs), carbon monoxide (CO), and coarse (PM_10_) and fine (PM_2.5_) particulate matter — all necessary for the atmospheric composition calculations — were produced for the target years on a nominal 1km x 1km grid covering the United Kingdom. Emissions data were internally re-projected and processed by the EMEMP4UK model to provide output concentration data at the model grid resolution of 0.055° x 0.055° resolution (∼5km x 6km) for the UK. The concentrations of PM_2.5_ calculated by the EMEP4UK model were used in conjunction with the residential history data (as described above). The sources of primary emitted PM are varied but the main contributors are essentially fuel combustion (from all sources) and the use of any mobile machinery, including road traffic. This is in contrast to secondary produced PM — such as ammonium sulphate which is formed by the interaction of ammonia gas and sulphur dioxide — which are strongly linked to specific sectors, such as SO_x_ (energy) and NH_3_ (agriculture). The PM components included in the EMEP4UK model are; primary PM, secondary inorganic and organic aerosols, sea salt, and mineral dust.[22] Although this work is focused on the UK the EMEP4UK requires emission data for the whole of Europe to account for the transboundary imports/export. EU data were kindly supplied at a 50km x 50km resolution[28, 29]. The EMEP4UK model is driven by 3D hourly meteorology calculated by the weather and research forecast model.[30] The meteorological year used for the 1935 and 1950 emission scenario was the year 2014, for the 1970, 1980, 1990 emission scenario was the year 2012, and for the 2001 emission scenario the year was 2001.

For the years 1970, 1980 and 1990, emission data in the official UK inventory, the National Atmospheric Emissions Inventory[31], were used to scale 2017 spatial distributions (1km x 1km resolution) of sectoral totals per pollutant, reported via the Selected Nomenclature for sources of Air Pollution system (SNAP sectors). While the use of contemporary distributions back to 1970 is imperfect, the majority of the time series had the best possible emissions estimates per sector. Emissions for 1950 were estimated and distributed in the Long Term Large Scale project[32], while the 1935 emissions were a scaled version of the 1950 distributions based upon activity data research, using the same spatial methods. Non-NH_3_ activity data prior to 1970 are largely a reflection of the use of fossil fuels such as coal and of oil-derived products such as diesel (DERV); coal usage in the UK had a double peak either side of World War Two before a rapid decline in the 1960s. Agricultural activity data such as animal numbers, principally associated with emissions of NH_3_, were derived from the Vision of Britain database.[33] For source strength emission factors (EFs), many were similar to those used by the UK National Atmospheric Emission Inventory (NAEI) in 1970 while in terms of the spatial distribution of pre-1970 data, the principal differences from the NAEI distributions were: power stations relevant to the time period were mapped and the distribution of industrial activity was tied to census employment data[32].

Raster files for each location year (1935, 1950, 1970, 1980, 1990, and 2001) were read into the **R** statistical computing environment version 4.0.2 using the raster package[34]. The latitude and longitude for each location were used to derive values from these raster files for each participant at each time point. Since participants may have had multiple locations within each time band, the unweighted mean of these multiple values per time point was calculated and used in the analyses as the value for that participant at that time point. For example, the mean of all the air pollution values corresponding to locations recorded between the years 1995 and 2004 would be allocated to the 2001 time point for an individual participant. For the purposes of sensitivity analyses, we also computed the maximum value for each time point per participant and the 90% percentile value.

### Statistical modelling

Following the convention in previous LBC1936 analyses, we modelled change in IQ score from age 11 to age 70 years separately from changes between the ages of 70, 76, and 79 years. The former used a linear regression model of *in utero* air pollution exposure (i.e., using PM_2.5_ data for 1935; this was the only measurement of air pollution which predated the MHT administration at age 11 years) and residualised change in IQ score from age 11 to age 70 years in the **R** statistical computing environment version 4.0.2. We adjusted this model for sex, parental occupation, and smoking status.

To estimate linear late life cognitive trajectories, we fitted latent growth models to IQ scores from ages 70, 76, and 79 years to estimate the average population cognitive curves. These longitudinal models permit estimation of the outcome’s mean and individual trajectories while permitting the inclusion of predictors (time-invariant or time-varying) to study their association with curve parameters such as the intercept and slope parameters. Latent growth models were estimated using maximum likelihood under a ‘missing at random’ missing data assumption. All latent growth models were estimated using MPLUS[35].

We summarise the models fitted in **Figure 1**, in which observed data are represented within rectangles, and latent variables such as the model’s intercept and slope are represented within circles. The one-way arrows indicate that the variable at the end of the arrow is explained in the model by the variable at the beginning of the arrow. Often in such figures, two-way curved arrows indicate covariances but as is commonly done – in order to simplify the figure – we have omitted these arrows here as well as the arrows that indicate error terms.

**Figure 1.**
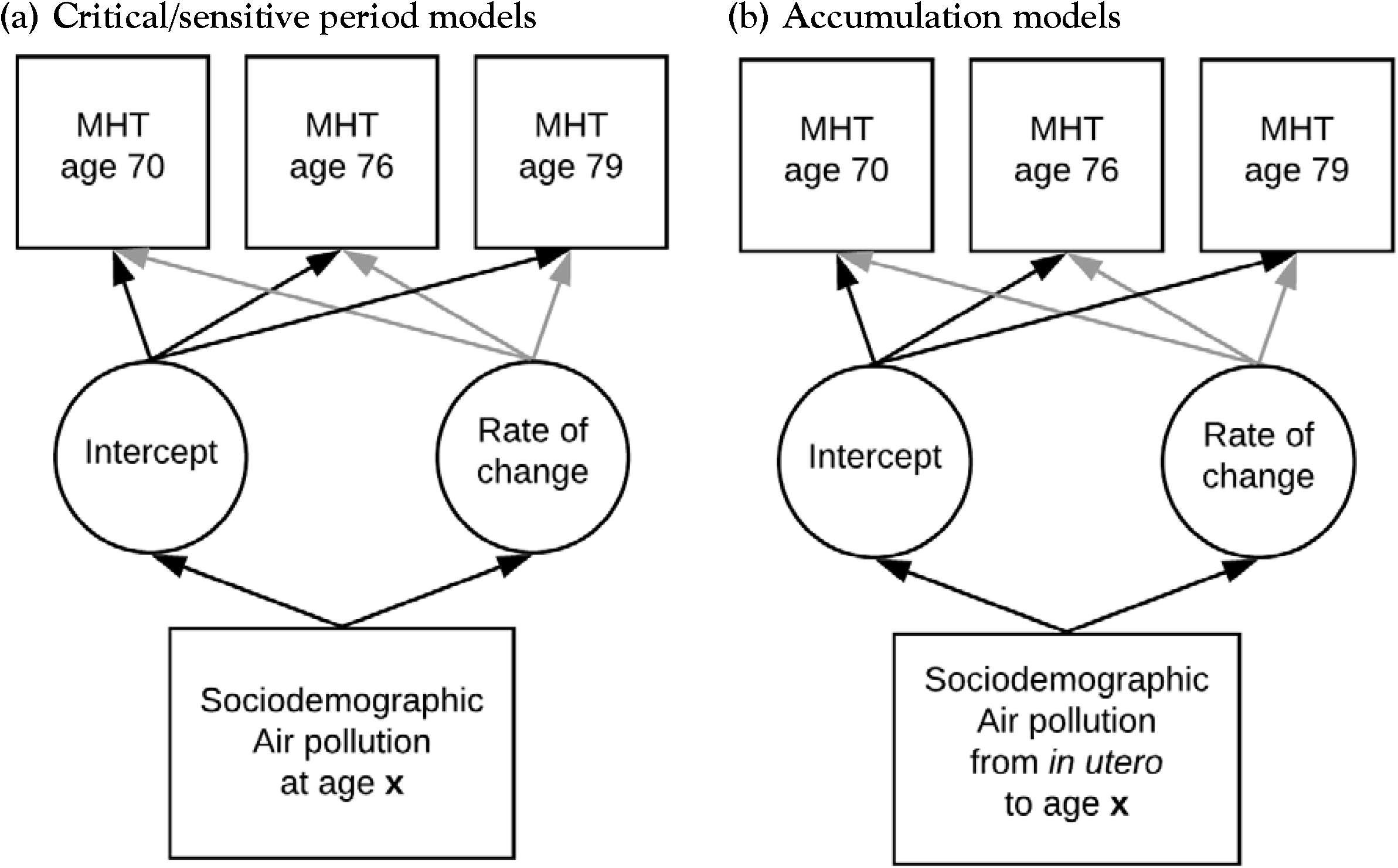
Figure representing the (a) critical/sensitive period and (b) accumulation models fitted to IQ scores: life course air pollution exposure and cognitive decline in the LBC1936

#### Life course models

The two main models in life course epidemiology are critical/sensitive periods and accumulation of risk[2]. To evaluate questions about any critical/sensitive period(s) of air pollution exposure and maximise the use of pollution data collected over the life course, we estimated late life trajectories of IQ scores at ages 70, 76 and 79 years, adjusting the intercept and rate of change for air pollution measures collected at different ages in the life course, age 11 IQ scores, sex, parental social class, and smoking status. Specifically, we adjusted the models separately for PM_2.5_ in 1935, 1950, 1970, 1980, 1990, or 2001. **Figure 1a** depicts an example of the critical/sensitive period model fitted here.

In order to evaluate an *accumulation of risk* model, we estimated a series of latent growth models similar to the previous ones, adjusting the level and rate of change for variables defined as the sum of air pollution to which the individual was exposed up to different stages in life. That is, we added *in utero* air pollution measures (i.e., from 1935) to air pollution measures collected in 1950 to derive an indicator of early life exposure; the sum of air pollution measures from 1935 to 1970 covered early life to young adulthood; additionally adding pollution from 1980 encompassed early life to mid-adulthood; the addition of air pollution in 1990 covered early life to late adulthood; finally, adding air pollution from 2001 covered early life to later life. **Figure 1b** depicts an example of the accumulation period model fitted here. **Text Box 1** summarises the models used in the present analyses.

## RESULTS

A total of 572 LBC1936 participants were included in the present analyses. Their characteristics are summarised in **Table 1**. Briefly, just under half were female, and had completed more than compulsory education. Just over a quarter had parents from occupational social classes I or II (i.e., less deprived), and about half were smokers at the time of recruitment to the LBC1936. Comparing the 572 LBC1936 participants for whom we had location (and therefore air pollution) data with the 519 participants excluded from these analyses revealed no major differences.

**Table 1.** Sample characteristics: life course air pollution exposure and cognitive decline in the LBC1936

|  | Included<sup>a</sup> | Excluded<sup>b</sup> | P<sup>c</sup> | Total LBC1936 sample |
| --- | --- | --- | --- | --- |
| N | 572 | 519 |  | 1091 |
| Age at SMS1947 (mean [SD] years) | 10.92 (0.27) | 10.96 (0.29) | 0.027 | 10.94 (0.28) |
| Female (%) | 46.9 | 53.0 | 0.0497 | 49.8 |
| Age 11 IQ<sup>d</sup> (mean [SD]) | 101.6 (15.0) | 98.2 (14.9) | <0.001 | 100.0 (15.0) |
| Parental occupational social class (% class I or II) | 27.7 | 26.3 | 0.011 | 27.1 |
| Current smoker at baseline (%) | 49.3 | 42.2 | 0.022 | 45.9 |

<sup>a</sup> Participants were included if they had at least one location recorded for each time period.
<sup>b</sup> Excluded participants included 21 with missing location data for at least one time period, 111 who did not respond to the questionnaire requesting lifetime residential history, and 387 who were not approached, mainly because they had died or withdrawn from the study prior to the questionnaire being used in 2014.
<sup>c</sup> P-values from comparisons of included and excluded participants
<sup>d</sup> 31 participants were missing age 11 intelligence data
<sup>e</sup> Self-reported
LBC1936: Lothian Birth Cohort 1936 (N=1091);
SMS1947: Scottish Mental Survey 1947 (N=70,805, of which the LBC1936 is a subset)

### Air pollution

**Table 2** shows the average air pollution estimates for the LBC1936 participants and **Supplementary Figure 1** shows the distribution of air pollution exposure at each time period. Supplementary Figure 2 shows participants’ PM_2.5_ exposure changes over time and Supplementary Table 1 shows the correlations between individuals’ PM_2.5_ exposure ranking at different time points. Rankings varied over time — likely due more to participants moving than the relative ranking of areas changing — but were more closely correlated when closer in time, suggesting it is feasible to explore critical/sensitive time periods. **Figure 2** shows the modelled PM_2.5_ values for Scotland in 1935; the urban centres are clearly visible.

**Table 2.** Annual average particulate matter (PM_2.5_) values at different time points for all participants: life course air pollution exposure and cognitive decline in the LBC1936

| **Year** | **Mean (sd)** | **Range** | **N<sub>total</sub><sup>a</sup>** | **>10µg/m<sup>3</sup><sup>b</sup>** |
| --- | --- | --- | --- | --- |
| 1935 | 34.8 (16.0) | 5.2-133.0 | 590 | 562 (95%) |
| 1950 | 32.4 (12.8) | 6.0-113.3 | 591 | 578 (98%) |
| 1970 | 17.0 (1.5) | 9.5-23.9 | 585 | 584 (100%) |
| 1980 | 15.0 (1.5) | 7.3-24.0 | 580 | 575 (99%) |
| 1990 | 13.4 (1.2) | 6.7-21.4 | 580 | 579 (100%) |
| 2001 | 7.9 (0.6) | 4.8-15.9 | 591 | 4 (0.7%) |

<sup>a</sup> 593 participants provided lifetime residential histories; 572 had air pollution data from all time periods and were included in the present analyses
<sup>b</sup> The number (%) of participants whose PM<sub>2.5</sub> exposure exceeded the WHO guidelines of an annual mean of ≤10µg/m<sup>3</sup>

**Figure 2.**
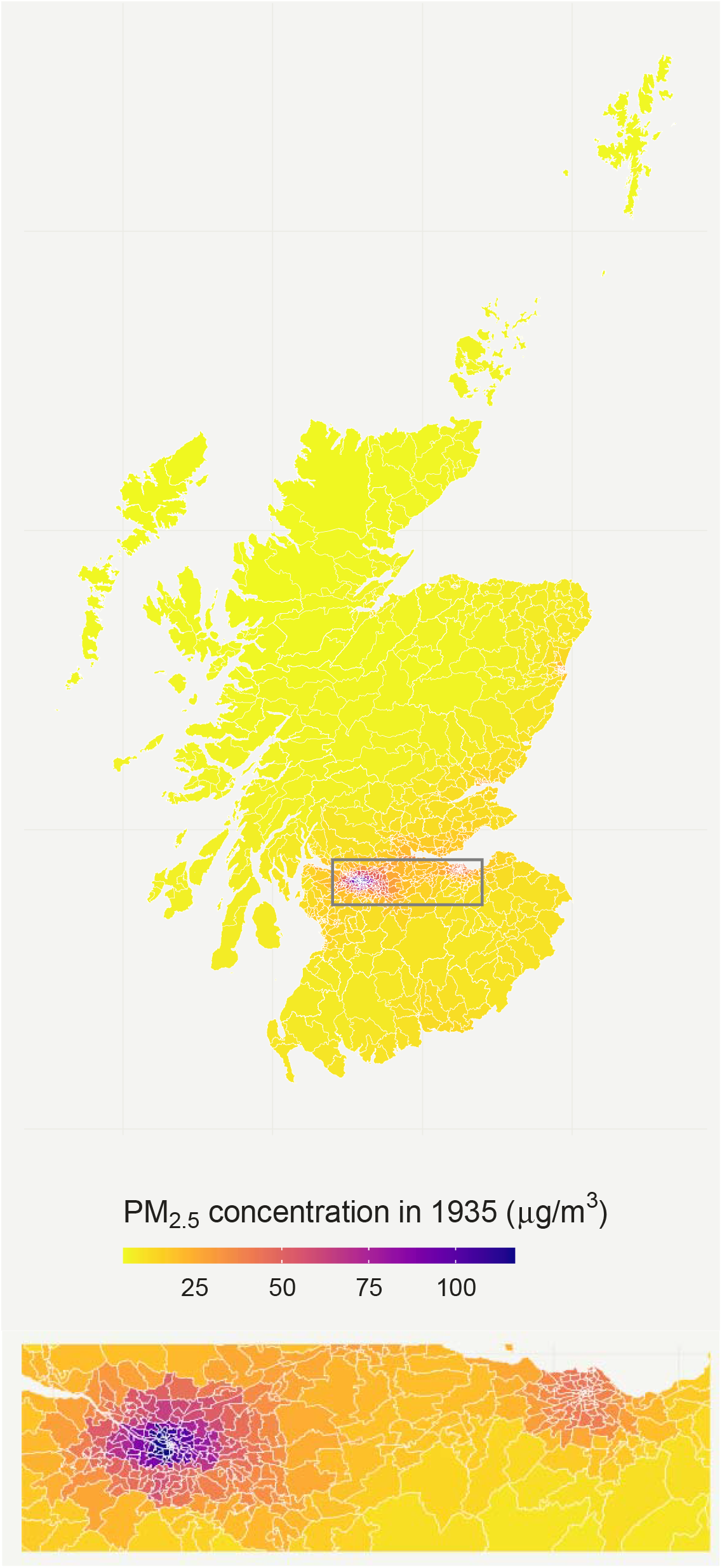
Modelled particulate matter (PM_2.5_) values in 1935: life course air pollution exposure and cognitive decline in the LBC1936 The area displayed in the lower panel and enclosed in a box on the upper panel is the central belt of Scotland including Glasgow (left) and Edinburgh (right). Over half of the population of Scotland lives in this area.

#### Is in utero air pollution exposure associated with cognitive trajectories over the life course?

Results from this model are presented in Table 3a, where the potential impact of air pollution measured in 1935 on the residualised change in IQ between the ages of 11 and 70 years, controlling for sex, parental social, class, and smoking was explored. There was a small association between higher levels of air pollution exposure in 1935 and *in utero* and a poorer IQ trajectory in IQ from 11 to 70 years (β=-0.006 IQ point per 1 μg/m^3^ increase in PM_2.5_, SE=0.002, P=0.03).

**Table 3.**
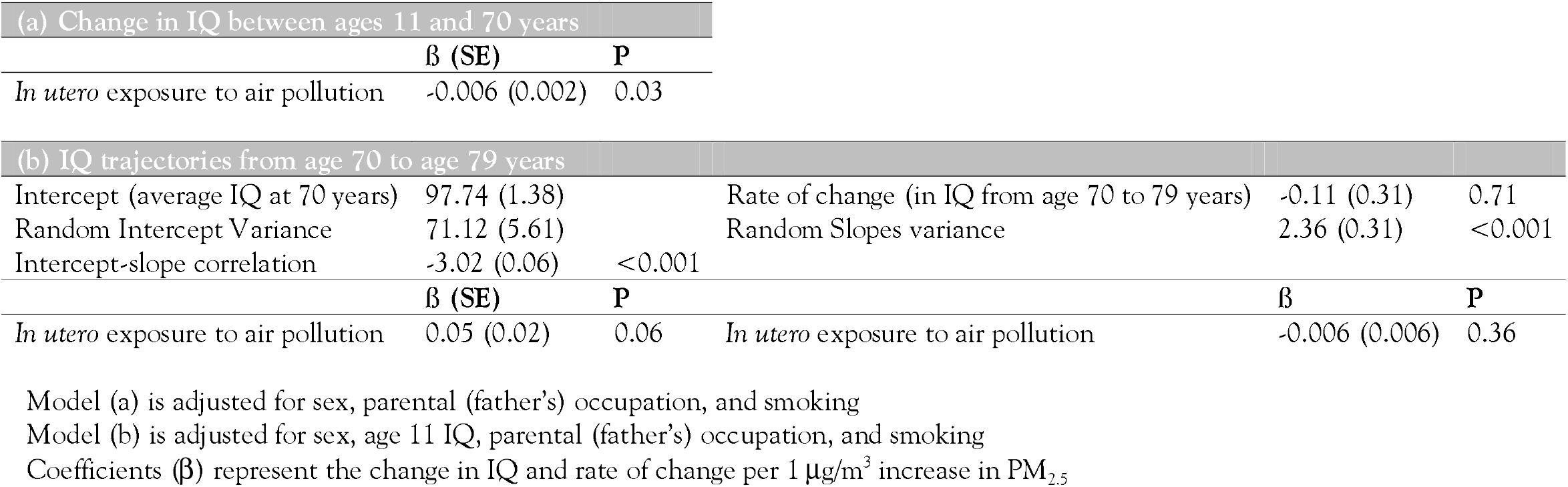
Results from (a) linear regression of residualised change in IQ from age 11 to age 70 years and (b) latent growth models fitted to IQ scores to estimate cognitive trajectories at ages 70, 76, and 79 years: life course air pollution exposure and cognitive decline in the LBC1936

#### Is in utero exposure to air pollution associated with late life cognitive trajectories?

**Figure 1a** depicts the model that estimates linear changes in IQ scores between the ages of 70, 76, and 79 years. In this model, the intercept represents the average IQ score at age 70 for a reference individual (a male whose father had a skilled job, who was exposed *in utero* to average levels of pollution, and who had an average IQ score at age 11 years) and the slope, the average rate of change of IQ scores from age 70 to 79 years. There was a small association between air pollution values for 1935 and the intercept (IQ score at age 70 years), albeit only of marginal statistical significance at conventional levels (P=0.06), but no association with rate of change in IQ score from age 70 to 79 years (P=0.36; Table 3b).

#### Critical/sensitive period

For the sake of brevity, Table 4 only contains estimates of the average value of IQ scores at age 70, their rate of decline until age 79 (for a reference individual, as defined above) and estimates of the association of air pollution exposure at each of the life course time points (apart from 1935 which was reported above) with IQ level at age 70 and rate of change (Figure 1b). No period of air pollution exposure had an effect on either the intercept or the rate of change of the models which reached statistical significance at conventional levels (all P>0.05).

**Table 4.**
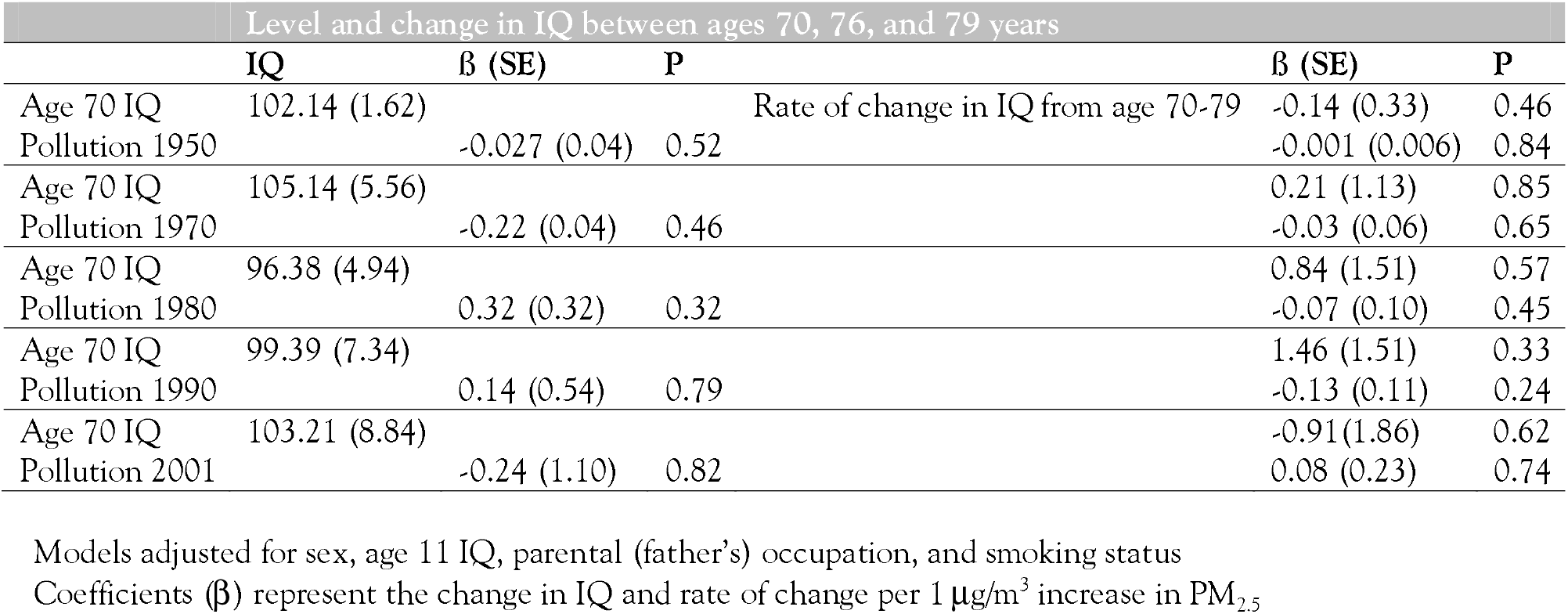
Estimates of the association between air pollution exposures at different time points in the life course with mean IQ at age 70 and its rate of change from 70 to 79 years: life course air pollution exposure and cognitive decline in the LBC1936

#### Accumulation model

Summary results of the models are presented in **Table 5**, where estimates of average IQ level at age 70, average IQ linear rate of change from that same age, and coefficients of the association between air pollution exposure at different stages of life with these parameters are presented. None of the risk periods had an effect on either the intercept or the rate of change of the models which reached statistical significance at conventional levels (all P>0.05). Our findings were robust to the sensitivity analyses varying the aggregation method used for multiple air pollution values.

**Table 5.**
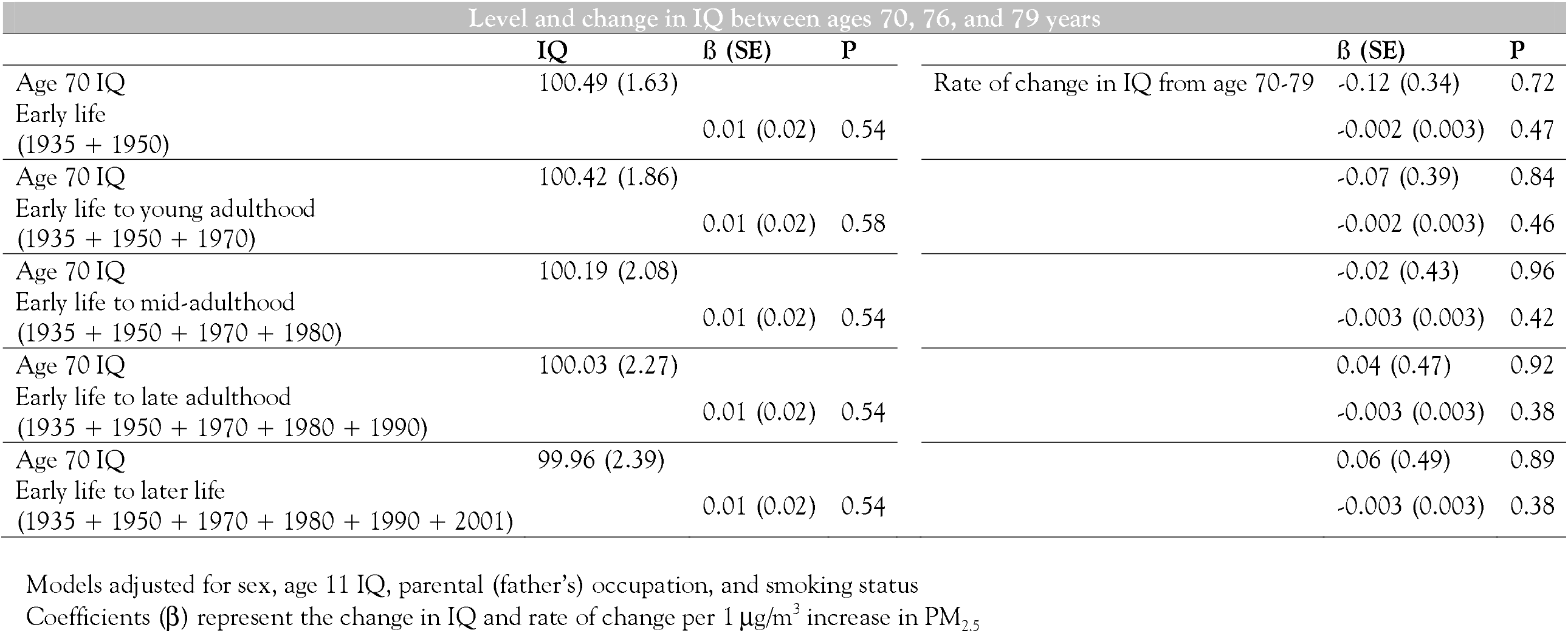
Estimates of IQ intercept (at age 70 years) and rate of change from age 70 and of the association of cumulative air pollution exposure at various stages of life: life course air pollution exposure and cognitive decline in the LBC1936

## DISCUSSION

Our main finding is that it is feasible to model historical air pollutant concentration data and incorporate them in epidemiological models to explore the influence of exposure to air pollution across the life course. We found little evidence that exposure to air pollution at different stages of the life course was associated with cognitive health and there was no support for an accumulation of risk. There was some evidence of exposure to air pollution *in utero* being associated with worse cognitive change between the ages of 11 and 70 years, but the effect size was small (β=-0.006). In particular we would highlight that these results have large degrees of uncertainties, considering the various methodologies used to produce the different air pollution concentration estimates due the wide range of emissions estimates, particularly for earlier estimates which have a lack of measured air quality data against which to be evaluated.

### Comparison with other literature

As noted in the introduction, there is a growing wealth of literature on the association between air pollution and subsequent cognitive impairment and dementia, but the majority of publications share the same shortcomings[10]: (1) an inability to explore *when* in the life course exposure to air pollution has the most impact?; (2) which pollutant(s) or components are most important?; and (3) since dementia describes a heterogeneous group of conditions, which are most affected by exposure to air pollution?

Investigators from the Washington Heights–Inwood Community Aging Project (WHICAP) and the Northern Manhattan Study (NOMAS) recently reported their findings of the impact of “long-term” exposure to air pollution (nitrogen dioxide, PM_10_, and PM_2.5_), but these participants (aged 65 years or older) were only recruited in the early 1990s and only their residential address at the time of recruitment was used to estimate their exposure to air pollution; air pollution values for the year before recruitment were used as the exposure[11, 12]. Our study was able to track migration and movement throughout the life course, combined with modelled atmospheric concentration data covering most of the twentieth century, to give a much better estimate of each person’s exposure to air pollution at different points in their lives.

### Limitations and Strengths

Referring to the three criticisms of the air pollution literature described above,[10] the present study could potentially shed some provisional light on the first (when in the life course is most important), but not the second or third. A decision was taken early on to minimise the impact of multiple testing by restricting the pilot modelling (of 1935 data) to a single pollutant; PM_2.5_ was chosen since the majority of the literature linking air pollution and dementia focused on that pollutant. Modelling other pollutants — such as smaller PM, NO_x_ etc. — is feasible and we hope to do this in the future, now that the feasibility of this approach has been demonstrated.

The collection of lifetime residential histories is rare and greatly augments the other data available in the LBC1936. However, there are limitations to the approach taken (retrospective collection of residential address history), including the fact that it is prone to recall bias. Furthermore, only participants who were alive in 2014 were approached, additionally introducing survivor bias. Finally, the accumulated PM_2.5_ exposure was calculated using an unweighted method – i.e. not taking into account the length of time an individual lived at each address. Our main aim in the present analysis was to establish proof of concept and hope that a more sophisticated weighted calculation – which could arguably be more accurate – could be taken by future studies.

Almost all participants — until 2001 — were exposed to levels of PM_2.5_ in excess of the World Health Organization’s guidelines of a maximum annual mean of 10μg/m^3^[36]. For comparison, approximately half of UK Biobank participants were exposed to similarly excessive values at baseline (mean [SD] 10.0 [1.1])[37], in line with most of the world[38]. There was a general reduction in air pollution over time — and marked step-changes between some time points — but it is unclear how much of this is artefactual, relating to methodological differences between the procedures used to generate these historical estimates. There are inherent uncertainties at all stages of the emissions estimation process, even for the present day, and many of these problems are magnified when trying to recreate an historical context. The lack of measurement data to verify source strength, the lack of data regarding the chemical composition of fuel and the behaviour of combustion technologies prior to emissions mitigations are just some of the many issues that can influence the uncertainty. From 1970 onwards, emissions uncertainties were estimated from the UK Inventory[39] while for 1935 and 1950, uncertainty estimations are expert judgement.[40] Given the assumptions that the fuel use data are lacking some detail (such as certain oil-based products, wood etc.), combustion technology was a lot more polluting than in 1970 due to a lack of various mitigating options such as scrubbers and particulate filters and that there are some missing sources such as construction etc., it is very likely that the uncertainty range is asymmetric with respect to the best estimate. To reflect this probable under estimation, the asymmetry was estimated to be one order of magnitude centred on the mean, that is 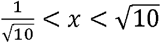 example, the emissions of PM_2.5_ in 1935 were 715kt (range 226-2261kt). Whilst the uncertainties were not utilised within the EMEP4UK model, it is important to note these qualitative estimates and the potential impacts on the final results. **Figure 3** shows the final emissions estimates for all pollutants per year (2015 is displayed for context).

**Figure 3.**
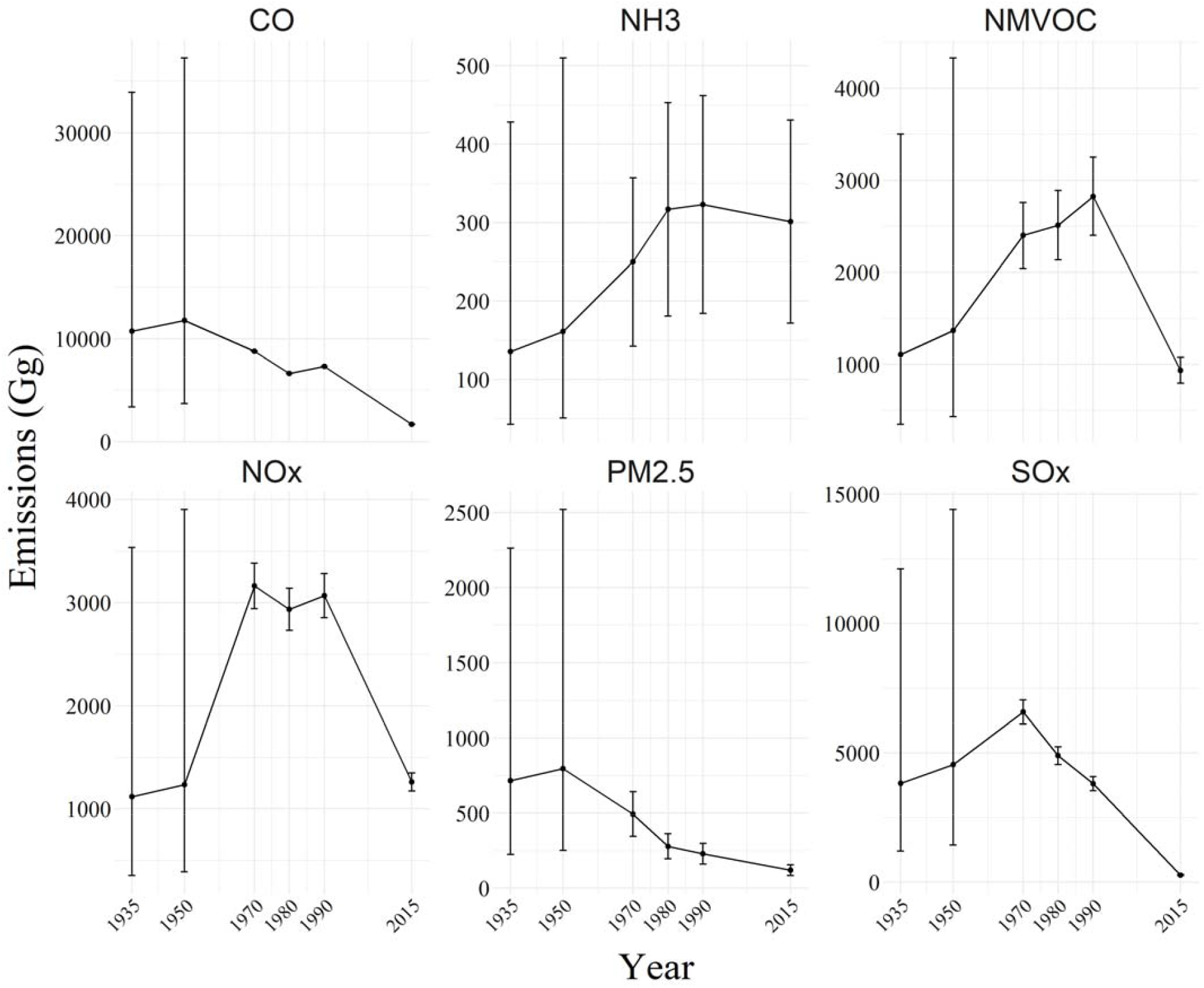
Modelled emissions totals (Gg) with uncertainty ranges for five air pollutants (CO, NH_3_, NMVOCs, NO_x_, and SO_x_), plus PM_2.5_, across five model years (2015 is included for context) for use in the EMEP4UK model: life course air pollution exposure and cognitive decline in the LBC1936

The focus of this paper is cognitive change rather than dementia. It is important to assess pre-dementia cognitive change and its determinants in their own right, but dementia is inarguably important. However, there were not sufficient LBC1936 participants who had developed dementia to allow meaningful models to be constructed. A comprehensive programme of dementia ascertainment in LBC1936 participants (who are currently in their mid-80s) is currently underway and, once these data are available, similar models focused on dementia will be possible.

Statistically, the life course model (change between ages 11 and 70 years) and late life models (intercept and change from 70 to 79 years) are not comparable. Furthermore, a linear assumption for the late life cognitive trajectories may be too strong. It may be necessary to explore more complex models, such as quadratic trajectories, but the primary aim of this study was proof of concept and so we have not taken that approach here. With additional time points, we could have constructed a model that would permit estimation of piecewise trajectories. This may become possible as further waves of data become available — wave 5 of the LBC1936 was completed last year and wave 6 was due to begin in Spring 2020 but had to be delayed because of the Covid-19 pandemic.

### Future directions

This paper is the first step towards an understanding of the associations between air pollution and cognitive decline and dementia from a life course epidemiology perspective. The modelled historical air pollution data need to be refined and harmonised across different time points – and these data used to provide a robust estimate of life course exposure – but we believe that we have demonstrated the feasibility and value of this approach. However, these air pollution data will be of little value without well-characterised cohort studies with full residential histories for participants, such as are available for the LBC1936. All high quality longitudinal cohort studies should explore whether it is possible to obtain such data through record linkage or — as in the LBC1936 — self-report.

### Conclusions

We have shown the feasibility of modelling historical air pollution data and incorporating them in epidemiological models. This is the first step in a new area and we look forward to a greater understanding of the life course effects of air pollution on the brain in coming years.

## Data Availability

Data are available to bona fide research collaborators subject to the standard agreements required by the LBC1936 study.

## ACKNOWLEDGEMENTS

This study was funded by the “Improving Health with Environmental Data” call from the Natural Environment Research Council, the Chief Scientist Office, and the Medical Research Council (NE/P010849/1). It follows research undertaken under the Mobility, Mood, and Place research project (2013–2016), supported by Research Councils UK (EP/K037404/1) as part of the Cross-Council Lifelong Health and Wellbeing Program under Principal Investigator Catharine Ward Thompson and Co-Investigators Jamie Pearce and Niamh Shortt.

TCR and the late JMS are/were members of the Alzheimer Scotland Dementia Research Centre funded by Alzheimer Scotland. The Lothian Birth Cohort 1936 is funded by Age UK (Disconnected Mind grant). TCR, the late JMS, and IJD were members of the University of Edinburgh Centre for Cognitive Ageing & Cognitive Epidemiology (which recently closed), part of the cross council Lifelong Health and Wellbeing Initiative (MR/K026992/1). Funding from the Biotechnology and Biological Sciences Research Council and Medical Research Council is gratefully acknowledged for the latter.

Initial findings of this study were presented at the Alzheimer’s Association International Conference in Chicago, 2018 (https://doi.org/10.1016/j.jalz.2018.06.2861).

---

**Text Box.** Summary of the models used in the present analyses

| OUTCOME | EXPOSURE |
| --- | --- |
| **Sensitive/critical period(s)** |  |
| Change in IQ from age 11 to age 70 years | In utero PM<sub>2.5</sub> exposure (1935) |
| Trajectories of IQ from age 70 to 79 years<br>(intercept and rate of change) | *In utero* PM<sub>2.5</sub> exposure (1935)<br>PM<sub>2.5</sub> exposure aged ~14 years (1950)<br>PM<sub>2.5</sub> exposure aged ~34 years (1970)<br>PM<sub>2.5</sub> exposure aged ~44 years (1980)<br>PM<sub>2.5</sub> exposure aged ~54 years (1990)<br>PM<sub>2.5</sub> exposure aged ~65 years (2001) |
| **Accumulation of risk** |  |
| Trajectories of IQ from age 70 to 79 years<br>(intercept and rate of change) | Early life<br>(1935 + 1950)<br>Early life to young adulthood<br>(1935 + 1950 + 1970)<br>Early life to mid-adulthood<br>(1935 + 1950 + 1970 + 1980)<br>Early life to late adulthood<br>(1935 + 1950 + 1970 + 1980 + 1990)<br>Early life to later life<br>(1935 + 1950 + 1970 + 1980 + 1990 + 2001) |

**Supplementary Figure 1a.**
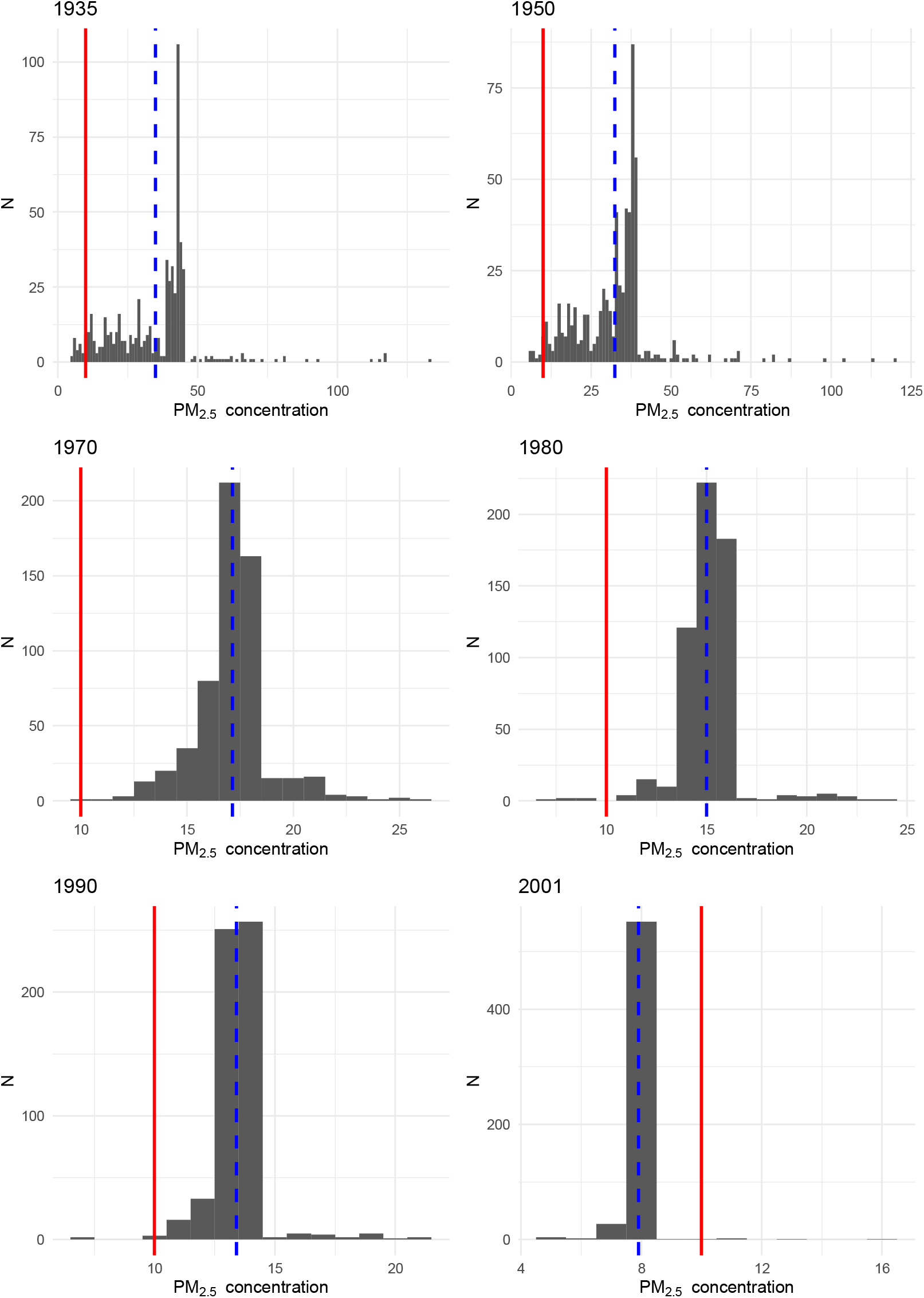
Mean PM_2.5_ exposure for each participant at each time point for which air pollution concentration data were modelled: life course air pollution exposure and cognitive decline in the LBC1936 Blue dotted line — mean PM_2.5_ value; Solid red line — WHO guidelines (annual average ≤10μg/m^3^)

**Supplementary Figure 1b.**
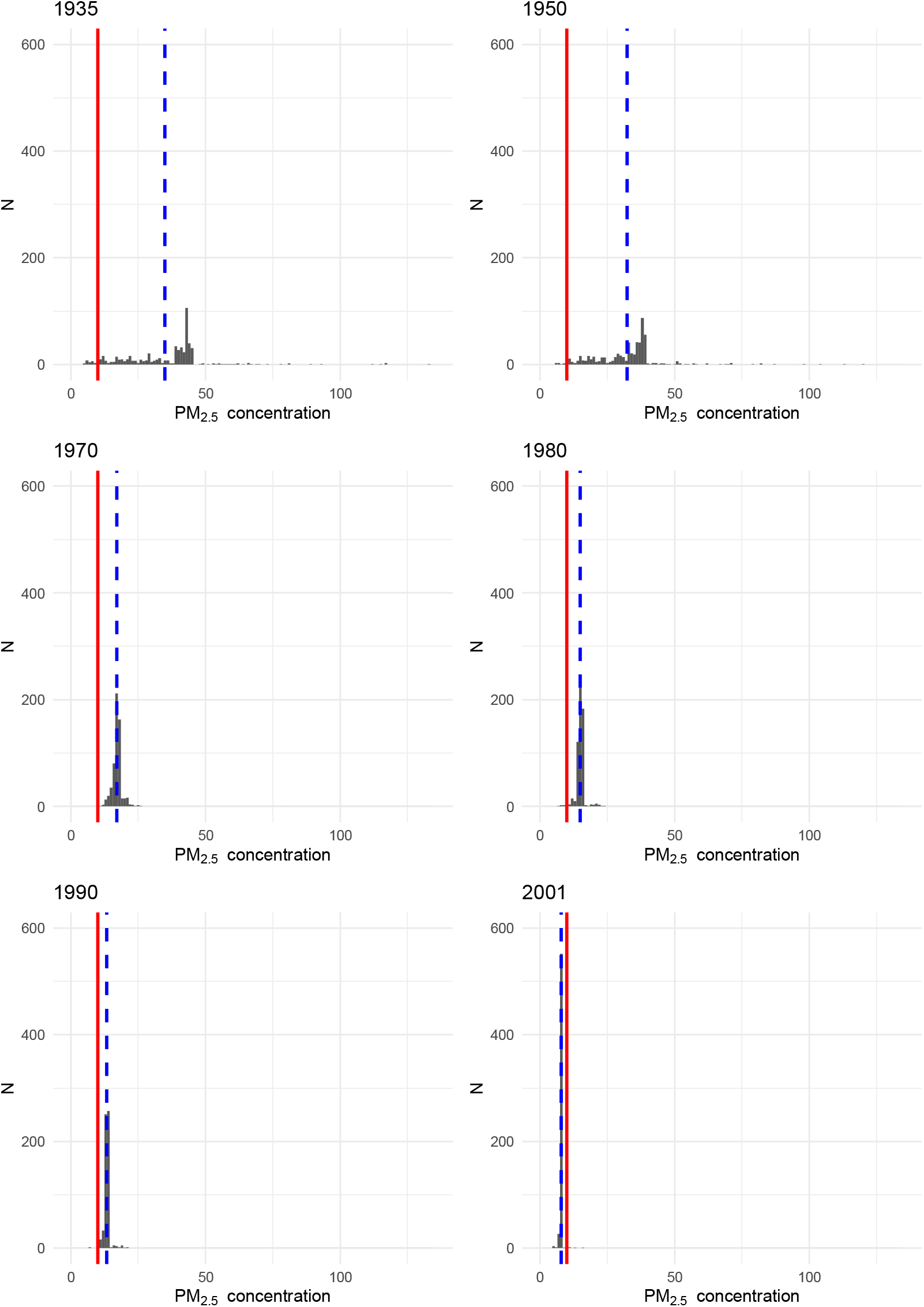
Mean PM_2.5_ exposure for each participant at each time point for which air pollution concentration data were modelled (all plotted on the same x and y scales): life course air pollution exposure and cognitive decline in the LBC1936 Blue dotted line — mean PM_2.5_ value; Solid red line — WHO guidelines (annual average ≤10μg/m^3^)

**Supplementary Figure 2.**
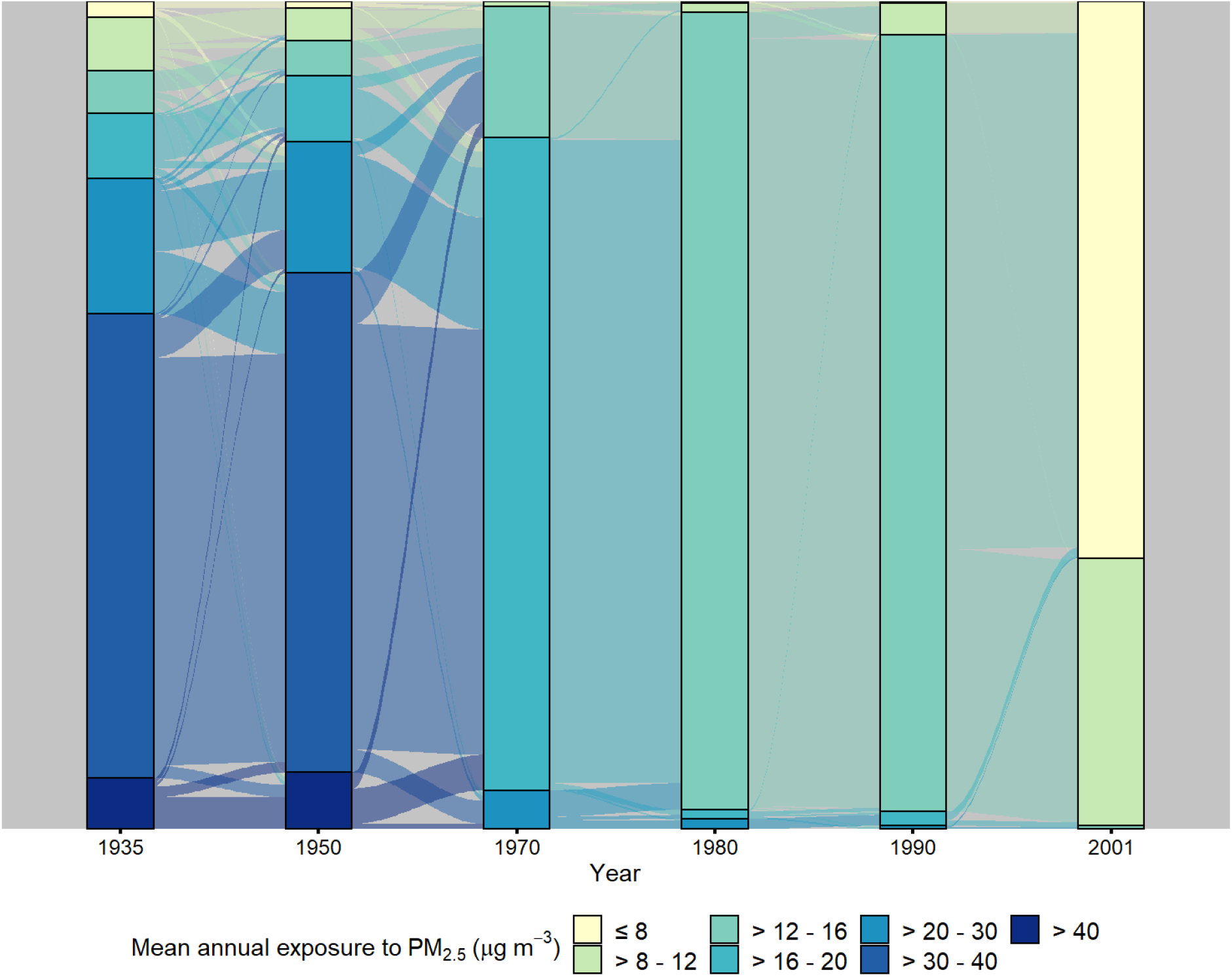
Sankey diagram indicating the change in mean annual PM_2.5_ exposure within individuals over time: life course air pollution exposure and cognitive decline in the LBC1936

**Supplementary Table 1.** Correlations between PM_2.5_ exposure rankings at different time points: life course air pollution exposure and cognitive decline in the LBC1936

|  | 1935 | 1950 | 1970 | 1980 | 1990 | 2001 |
| --- | --- | --- | --- | --- | --- | --- |
| 1950 | 0.53 | - |  |  |  |  |
| 1970 | 0.13 | 0.22 | - |  |  |  |
| 1980 | 0.06 | 0.18 | 0.57 | - |  |  |
| 1990 | 0.09 | 0.19 | 0.53 | 0.82 | - |  |
| 2001 | 0.003 | 0.10 | 0.05 | 0.03 | 0.08 | - |

